# Structural Barriers and Socioeconomic Determinants of HIV-Related Mortality in Children Under Five: Mixed-Methods Evidence from the Child Health and Mortality Prevention Surveillance Program, Sierra Leone

**DOI:** 10.64898/2026.11.23.26368716

**Authors:** Sabrina Clumeck, Carlos Roque Rojas, Dickens Kowuor, Hassan Sahid Sesay, Soter Ameh, Omar Galárraga, Ikechukwu Udo Ogbuanu

**Affiliations:** Brown University School of Public Health, Providence, Rhode Island, United States of America; Watson School of International and Public Affairs, Providence, Rhode Island, United States of America; Centre for Health Economics (CHE), University of York, York, United Kingdom; The Africa Research Collaborative (The ARC), on behalf of the Child Health and Mortality Prevention Surveillance, Freetown, Sierra Leone; FOCUS 1000, Freetown, Sierra Leone; Department of Community Medicine, Faculty of Health Sciences, University of Calabar, Nigeria; Department of Global Health and Population, Harvard T.H. Chan School of Public Health, Boston, Massachusetts, United States of America; Hubert Department of Global Health, Rollins School of Public Health, Emory University, Atlanta, GA, United States of America

**Keywords:** Mother-to-child transmission, HIV, PMTCT, Minimally invasive tissue sampling, Socio-economic status, Geographic access, Health system barriers, CHAMPS, Sierra Leone

## Abstract

**Objective:** Developing equitable prevention of mother-to-child transmission (PMTCT) policies requires local evidence, yet how socioeconomic, geographic, and health system factors jointly drive HIV-related child mortality remains underexplored in Sierra Leone. This study evaluated the association between socioeconomic status (SES), geographic access, and pediatric HIV-related mortality, while examining the system-level and sociocultural barriers driving missed PMTCT opportunities.

**Materials and Methods:** We used a mixed-methods design. Quantitative analyses drew on Child Health and Mortality Prevention Surveillance Sierra Leone surveillance data (2019–2024) for deceased under five years old (U5s) with confirmed HIV status. Stepwise logistic regression evaluated associations between covariates (SES, maternal education, distance to health facilities) and a composite binary outcome of HIV-related death. Qualitatively, 33 key informant interviews with PMTCT stakeholders were analyzed using thematic framework analysis in Dedoose.

**Results:** The quantitative sample included 1,230 deceased children, of whom 162 (13.2%) reported an HIV-related death. Maternal education and distance to health facilities were statistically significant predictors of HIV-related death, whereas wealth quintiles showed no association. The qualitative findings identified system-level constraints disrupting the PMTCT cascade, including delayed entry into antenatal care, weak postnatal follow-up, stockouts of HIV test kits and antiretroviral drugs, limited polymerase chain reaction capacity and delayed results, and gaps in scheduling, tracking, and data systems. Donor-driven targets and funding constraints shaped procurement and service availability. Stigma, fear of disclosure, confidentiality concerns, transport costs, and poverty further undermined engagement and retention.

**Conclusions:** Pediatric HIV infection and related outcomes were driven by cumulative barriers across access, service readiness, and sociocultural context rather than household wealth alone. Improving PMTCT outcomes requires strengthening diagnostic and supply chain infrastructure, enhancing patient-tracking systems, and delivering stigma-sensitive, community-anchored care.

## 1. Introduction

Mother-to-child transmission (MTCT) of HIV remains a major public health challenge in sub-Saharan Africa, including Sierra Leone [1]. Despite substantial global progress in implementing prevention of mother-to-child transmission (PMTCT) programs, these gains remain vulnerable to health system constraints and social vulnerabilities that limit equitable access to care [2,3]. The Joint United Nations Programme on HIV/AIDS (UNAIDS) and the World Health Organization (WHO) have established the 95-95-95 targets to eliminate HIV as a public health threat. However, Sierra Leone continues to fall short, particularly with respect to early diagnosis and timely treatment initiation among HIV-positive pregnant women. Low antenatal HIV screening rates lead to missed maternal diagnoses and missed opportunities to deliver PMTCT interventions, which can result in preventable pediatric HIV infections and AIDS-related child deaths; delayed diagnosis also postpones infant identification and timely ART initiation [2,4].

Breakdowns along the PMTCT continuum are driven in large part by fundamental operational constraints within the health system. Early infant diagnosis coverage in Sierra Leone remains critically low, with only 8.5% of HIV-exposed infants receiving testing within the first two months of life, a period essential for initiating life-saving ART. These missed diagnostic opportunities are further exacerbated by recurrent stockouts of HIV test kits and antiretroviral drugs, which delay treatment initiation for mothers and infants and disrupt continuity of care, as documented in both health system assessments and recent field reports [5–7]. Chronic shortages of trained health personnel, particularly in rural and under-resourced facilities, further constrain the consistent delivery of PMTCT services, contributing to loss to follow-up and avoidable infant deaths.

These operational failures reflect deeper structural weaknesses in procurement and financing systems. Fragmented supply chains, vertical donor programming, and prolonged external dependence have been shown to undermine coordination and efficiency within Sierra Leone’s health sector, weakening the reliability of essential commodity distribution [8]. The absence of effective pooled procurement mechanisms further inflates costs and undermines supply stability. While pooled procurement has demonstrably improved affordability and availability of HIV diagnostics and treatment in comparable settings, legal and institutional barriers have limited its adoption in Sierra Leone [9–11]. As a result, structural inefficiencies continue to translate directly into operational breakdowns across the PMTCT cascade.

Demand-side barriers further shape who can engage with an already fragile health system. HIV- related stigma remains a pervasive deterrent to antenatal attendance, HIV testing, and sustained ART adherence, discouraging timely engagement with PMTCT services [12,13]. In addition to stigma, poverty and economic vulnerability have long been recognized as important structural determinants of HIV transmission and barriers to effective prevention, particularly in low- and middle-income countries [14]. Socio-economic status (SES) also plays a decisive role in shaping HIV risk and health outcomes across sub-Saharan Africa [15,16]. Evidence from southern Mozambique demonstrates that individuals in the lowest wealth quintiles experience substantially higher odds of HIV infection compared to wealthier groups, with women disproportionately affected across all socio-economic strata [17]. However, evidence from the 2019 Sierra Leone Demographic and Health Survey indicates a different pattern: women in the middle, richer, and richest wealth quintiles showed significantly higher odds of HIV risk factors compared with those in the poorest quintile, highlighting the context-specific relationship between socio-economic status and HIV vulnerability in Sierra Leone [18]. Together, stigma and socio-economic disadvantage act as filters that determine who is able to access, navigate, and benefit from PMTCT services.

Technological and biomedical innovations offer clear opportunities to reduce MTCT, yet their potential remains unrealized in many low-resource settings. Decentralized early infant diagnosis (EID) using dried blood spot (DBS) testing has improved diagnostic coverage and reduced result turnaround times in other contexts, including Thailand [19]. Similarly, emerging biomedical strategies such as broadly neutralizing antibodies (bNAbs) and long-acting antiretroviral therapies, including lenacapavir, have the potential to improve adherence and reduce transmission risk, particularly where daily ART regimens are difficult to sustain [20]. However, access to these innovations in Sierra Leone remains constrained by high costs, procurement barriers, intellectual property restrictions, and limited delivery capacity [21–23]. This tension highlights a central challenge: effective tools exist, but health systems lack the capacity to deliver them equitably.

Understanding how health system weaknesses and socio-economic vulnerability intersect to shape HIV outcomes in children is therefore critical in the Sierra Leonean context. Locally grounded evidence is essential for developing PMTCT policies that advance both equity and effectiveness. However, the interaction between socio-economic disadvantage and systemic failure in shaping HIV-related child mortality remains insufficiently explored. To address this gap, this study has two objectives: first, to estimate the association between access determinants including SES and HIV-related mortality among deceased children enrolled in the Child Health and Mortality Prevention Surveillance (CHAMPS) program in Sierra Leone; and second, to explore the health system barriers undermining the effectiveness of PMTCT services, drawing on perspectives from frontline health workers and key stakeholders and examining operational challenges such as stockouts of HIV test kits, antiretroviral drug shortages, and delays in laboratory processing. By integrating quantitative mortality data with qualitative stakeholder insights, this study identifies critical system breakdowns along the PMTCT cascade and provides actionable evidence to guide equitable health system strengthening in Sierra Leone.

## 2. Materials and Methods

### 2.1 Study Design

This study employed a mixed-methods approach. The design for the quantitative component of the study was an analysis of a surveillance dataset, aimed at examining how access determinants, including SES, may be linked to HIV-related mortality among deceased children enrolled in the CHAMPS program in Sierra Leone. The qualitative component was a qualitative descriptive study aimed at exploring systemic barriers to the PMTCT of HIV. The mixed approach allows quantitative findings to be contextualized and explained using qualitative evidence, particularly in areas where surveillance data alone may not capture underlying mechanisms such as stigma, service disruptions, or care-seeking barriers.

### 2.2 Quantitative methods

#### 2.2.1. Data source

This study was conducted using data from the CHAMPS program in Sierra Leone. CHAMPS is a multi-country surveillance platform designed to generate data on causes of child mortality through enhanced post-mortem investigation and standardized data collection procedures [24].

Data were collected between 2019 and 2024 from defined catchment areas encompassing both urban and rural communities. These catchment areas are linked to sentinel health facilities and surrounding populations where child deaths are systematically identified and investigated. Data sources include clinical records, verbal autopsies, social autopsies, and laboratory findings, all harmonized within the CHAMPS data architecture. Briefly, CHAMPS procedures to confirm infections, including HIV, include the use of minimally invasive tissue sampling and a review by the Determination of Cause of Death (DeCoDe) panel of the clinical, laboratory and contextual data. Details of the procedures have been described previously [25] and are summarized in the Supplementary Text A.

#### 2.2.2 Study population

The study population was composed of stillbirths and deceased children under the age of five, from any cause, who were enrolled into the CHAMPS Sierra Leone study between 2019 and 2024. Inclusion criteria were the following:

- A confirmed cause of death determined through Minimally Invasive Tissue Sampling (MITS) and DeCoDe panel review
- A documented HIV status at or near the time of death, based on clinical abstraction or laboratory findings; or
- A DeCoDe diagnosis of sepsis or pneumonia as the underlying or immediate cause of death.

Children were excluded from the analysis if either cause of death determination or HIV status information was missing or indeterminate.

#### 2.2.3 Outcome

The main outcome is a binary indicator identifying deceased children with either confirmed HIV infection or any of the two intermediate conditions to HIV-related mortality: severe pneumonia or sepsis. This indicator is derived from data captured by the CHAMPS Child Clinical Data Abstraction tool. Cases are coded as ‘1’ if the child’s death was associated with either a laboratory- confirmed HIV infection or a qualifying intermediate condition, and zero otherwise. Outcome classification is based on a synthesis of available clinical records, laboratory results, and the standardized diagnostic review conducted as part of the CHAMPS post-mortem investigation protocol.

#### 2.2.4. Independent variables

Socio-economic status (SES), the primary exposure of interest in this analysis, is conceptualized as a multidimensional construct reflecting both household material resources and maternal human capital. Operationally, SES was measured using a composite household wealth index derived from data on asset ownership and infrastructure characteristics. These data were collected via the CHAMPS Social Autopsy instrument and encompassed indicators such as housing materials, sanitation facilities, access to electricity, and mobile phone ownership.

The wealth index was constructed following the established Demography and Health Survey (DHS) methodology, which employs Principal Component Analysis (PCA) (The DHS Program [Sierra Leone DHS], 2019). Although prevalent in public health research, the conventional DHS wealth index is critiqued for a systematic urban bias [26–29].

This bias arises because the first principal component is disproportionately influenced by asset patterns characteristic of urban areas (e.g., specific consumer durables). Consequently, wealth manifestations more common in rural settings—such as ownership of agricultural land or livestock—are often poorly captured, leading to a distorted representation of economic stratification in predominantly rural populations. The context of Sierra Leone exemplifies this limitation, where rural wealth and access to services are frequently intertwined.

To develop a more geographically neutral and context-sensitive measure, we augmented the traditional asset-based variables with metrics of access to key services and infrastructure, including distance to healthcare facilities. This adjustment aims to capture critical dimensions of economic opportunity and constraint relevant to both rural and urban households.

Finally, to integrate these diverse indicators coherently and maximize the index’s discriminatory power, we employed a polychoric dual-component PCA [30]. This technique is well-suited to mixed variable types (continuous and ordinal) and avoids the methodological shortcomings of using multiple binary dummy variables to represent a single latent asset construct. The resultant index provides a more robust and nuanced measurement of household wealth across the urban-rural continuum.

Multivariable models were adjusted for covariates selected based on theoretical relevance and data availability. These covariates are categorized into child-level, maternal, and health system factors. Child-level characteristics include age at death, categorized into clinically meaningful age groups (stillbirth, death in the first 24 hours, early neonate [1–6 days], late neonate [7–27 days], infant [28 days–11 months], and child [12–59 months]), and sex. Maternal characteristics include education (primary / secondary / higher) and religion (Christian / Muslim / other).

Health system and structural covariates were included to capture barriers to care not fully explained by individual or household characteristics. Geographic access to health services was treated as the primary structural exposure of interest. This was measured as distance to the nearest health facility.

Additional health system covariates included place of death (community or health facility) and place of residence (urban or rural), which were classified using land use and settlement layers derived from the Global Land Analysis and Discovery (GLAD) dataset [31] and accessed through national spatial datasets provided by the Sierra Leone Bureau of Statistics. Child residence points were spatially overlaid onto these polygons in QGIS (version 3.44) to determine classification. This variable accounts for structural disparities in infrastructure, service coverage, and population density that may confound the relationship between household socio-economic status and health outcomes. The method to measure geographic access can be found in Supplementary Text B.

Lastly, to capture additional structural determinants of health, we constructed and included a suite of geospatial and environmental covariates in the analysis. We obtained monthly climate data for the study period from the ERA5-Land reanalysis dataset [32]. For each child, mean monthly 2-meter air temperature and total monthly precipitation corresponding to the month and year of death were extracted at their residence coordinates using a Python-based raster extraction workflow (Python version 3.14.3), implemented with the Rasterio and GeoPandas libraries. Temperature values were converted from Kelvin to degrees Celsius and precipitation from meters to millimeters for analysis.

#### 2.2.5. Data analysis

The analysis proceeded in two distinct phases: (1) descriptive characterization of the study population, and (2) regression modeling to examine the relationship between access determinants including SES and the primary outcome.

Descriptive statistics were calculated to summarize the socio-demographic characteristics of the study population. Categorical variables are presented as frequencies and proportions, while continuous variables were summarized as means and standard deviations. Prior to regression modeling, missing data were assessed for all key variables. Observations with missing or indeterminate outcome status were excluded. For covariates, a complete-case analysis approach was employed. An exception was made for maternal education, where multiple imputation was used to address a substantial proportion of missing data given the variable’s theoretical relevance to the outcome.

To examine the association between access determinants, including SES and HIV, and intermediate outcome–related child mortality, we used a stepwise binary logistic regression approach. Initially, model 1 included wealth quintiles only and served as a baseline unadjusted assessment of socioeconomic gradients. Afterwards, model 2 adjusted for maternal education, place of residence, distance to the nearest health facility, and temperature at date of death. Lastly, model 3 further adjusted for child age group and sex to assess robustness and potential mediation. Model fit was evaluated using pseudo R-squared values and comparative likelihood ratio tests. Statistical significance was assessed using p-values derived from likelihood ratio chi-squared tests. Statistical analyses were performed using Stata (version 19; StataCorp LLC, College Station, TX, USA) [33].

### 2.3 Qualitative methods

#### 2.3.1. Data collection

Qualitative data were analysed using the Framework Method, a systematic approach to thematic analysis widely used in applied health research [25,34]. We carried out face-to-face key informant interviews with health workers and program stakeholders involved in the delivery, coordination, or oversight of PMTCT and HIV services in Sierra Leone. Participants were approached in person at their workplaces by the lead author, in collaboration with a national HIV program coordinator based in Makeni and Freetown. Individuals were identified based on their professional roles and direct involvement in maternal and child health or HIV service delivery.

Interviews were conducted at participants’ workplaces or in another quiet location of their choice, such as an office or meeting room, to protect privacy and minimize interruptions. Prior to each interview, participants received an information sheet, were given the opportunity to ask questions, and provided written informed consent. In line with standard CHAMPS practice, compensation was offered to acknowledge participants’ time rather than as an incentive to participate.

A total of 33 key informants were interviewed using purposive sampling. This sampling strategy ensured inclusion of participants with direct, practice-based experience of PMTCT implementation and health system processes across different service delivery settings. The sample included nurses, midwives, doctors, pharmacists, community health workers, NGO staff, program managers, and representatives of national and international HIV programs. Participants were recruited from two public hospitals, two private hospitals, several peripheral health units and clinics, the Ministry of Health, and HIV-focused implementing organizations.

Two researchers conducted the interviews: one led the interview, and the other took notes. Before the interview, participants completed a short self-administered questionnaire collecting socio-demographic information such as age, sex, years of professional experience, and current role. A semi-structured interview guide was developed by the lead author, informed by WHO and national PMTCT guidance, and reviewed with members of the SBSS (Social and Behavioral Sciences Systems) team and CHAMPS leadership to ensure contextual relevance. The guide was slightly adapted for different participant categories while maintaining a consistent core structure (Supplementary Text C).

All interviews were audio-recorded with participants’ permission, and the note-taker documented key observations, non-verbal cues, and contextual details. Interviews used open-ended questions with follow-up probes to encourage elaboration and clarification. Data collection continued until thematic saturation, defined as the point at which no substantively new information emerged across interviews and professional groups.

#### 2.3.2 Data analysis

All audio recordings were transcribed verbatim, and the lead author and members of the SBSS team checked the transcripts against the recordings and field notes to ensure accuracy and remove identifying information. We then used a thematic framework analysis approach to organise and interpret the material. The lead author and a qualitative researcher from *FOCUS 1000*, a Sierra Leone–based non-governmental organization focused on maternal and child health, first read all transcripts several times to become familiar with the data, note initial impressions, and identify recurring issues related to PMTCT service delivery, stockouts, laboratory systems, stigma, and access to care.

Transcripts and field notes were imported into Dedoose qualitative software for coding and data management. We coded the material line by line, combining deductive codes derived from the interview guide and the study objectives with inductive codes that emerged directly from participants’ narratives. Parent codes were used to capture broad themes such as “health system barriers,” “supply chain and stockouts,” “stigma and disclosure,” and “socioeconomic constraints,” while child codes captured more specific responses and examples within each area. Coding was led by the first author in close collaboration with the qualitative researcher; discrepancies in how codes were applied were discussed and resolved through consensus, and the coding framework was refined iteratively as new nuances appeared in the data.

After coding, we used Dedoose outputs and exported code-by-case matrices to Excel to help compare patterns across participant types and settings. Related codes were grouped into descriptive themes, which were then interpreted analytically in light of the study’s second objective: to explore health system barriers that may undermine the effectiveness of PMTCT services. The SBSS team reviewed the emerging themes and subthemes to assess credibility and ensure they reflected the range of perspectives shared in the interviews. Representative verbatim quotations were then selected to illustrate each main theme in the Results section.

To strengthen the trustworthiness of the analysis, several steps were taken. Credibility was supported through repeated reading of transcripts, triangulation between interview transcripts and field notes, and discussion of emerging findings with the SBSS team to ensure that interpretations reflected participants’ perspectives. Dependability was supported by using a structured coding process within Dedoose and maintaining a coding framework that was refined iteratively as the analysis progressed. Confirmability was strengthened through collaborative coding and consensus discussions between the lead author and the qualitative researcher, which helped reduce the influence of individual interpretation. Authenticity was supported by including verbatim quotations from participants in the Results section to illustrate key themes and represent the range of views shared during the interviews.

## 3. Results

### 3.1 Quantitative results

#### 3.1.1 Socio-demographic and clinical characteristics of study population

Table 1 presents the socio-demographic and clinical characteristics of the study population stratified by HIV-related death (yes/no). The analytic sample included 1,230 deceased children under five years of age, of whom 227 (18.5%) reported HIV-related death. More than half of the children were male (56.7%). HIV-related deaths were more frequent among males (61.1%) than females (38.9%). Age at death varied substantially across outcome groups. Stillbirths accounted for a smaller proportion of HIV-related deaths (2.8%) compared with other age groups, while infants and older children represented a larger share of outcome-positive deaths. Infants accounted for 32.1% of HIV-related deaths, compared with 15.5% among those without the outcome.

**Table 1.**
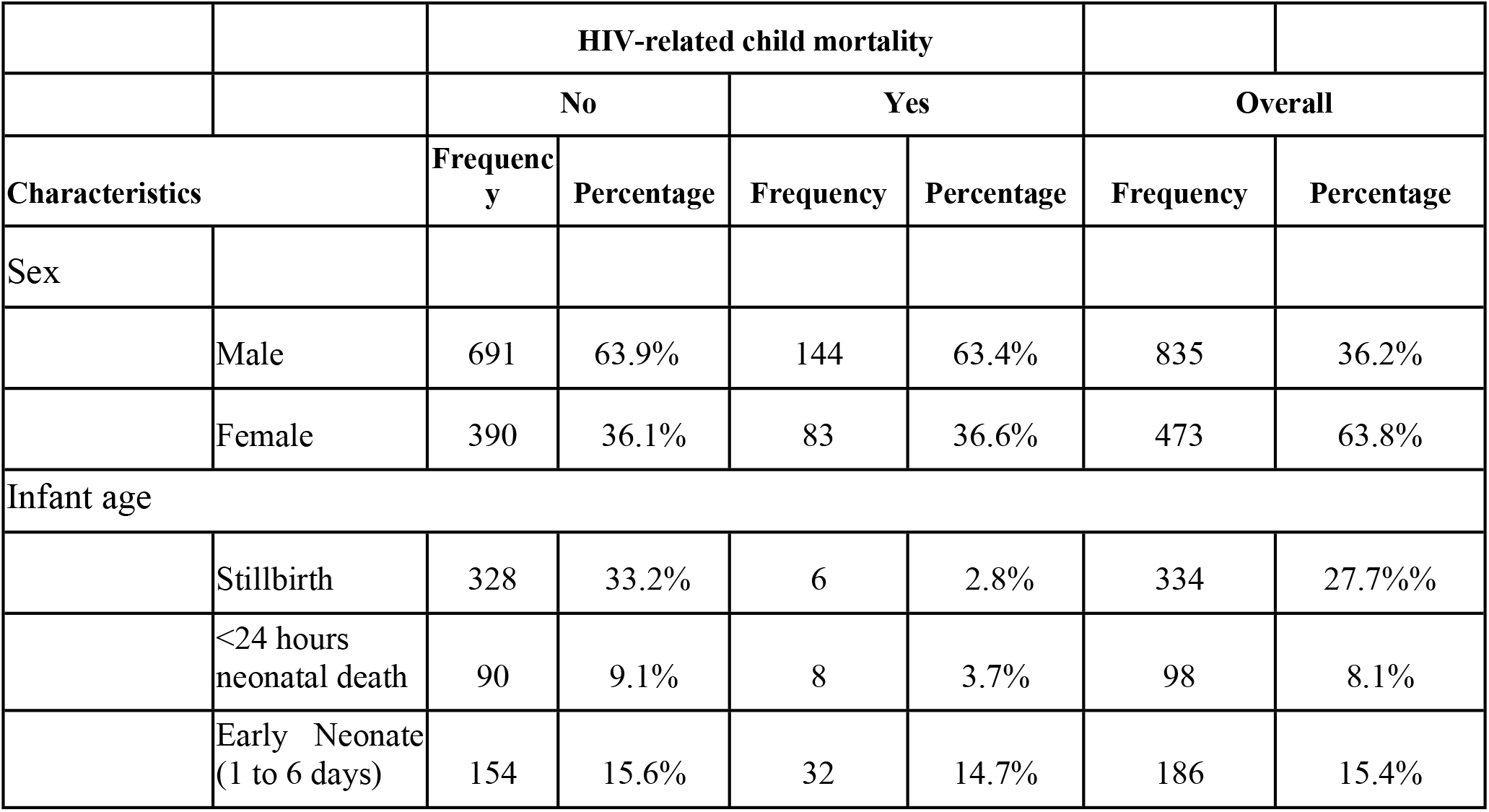
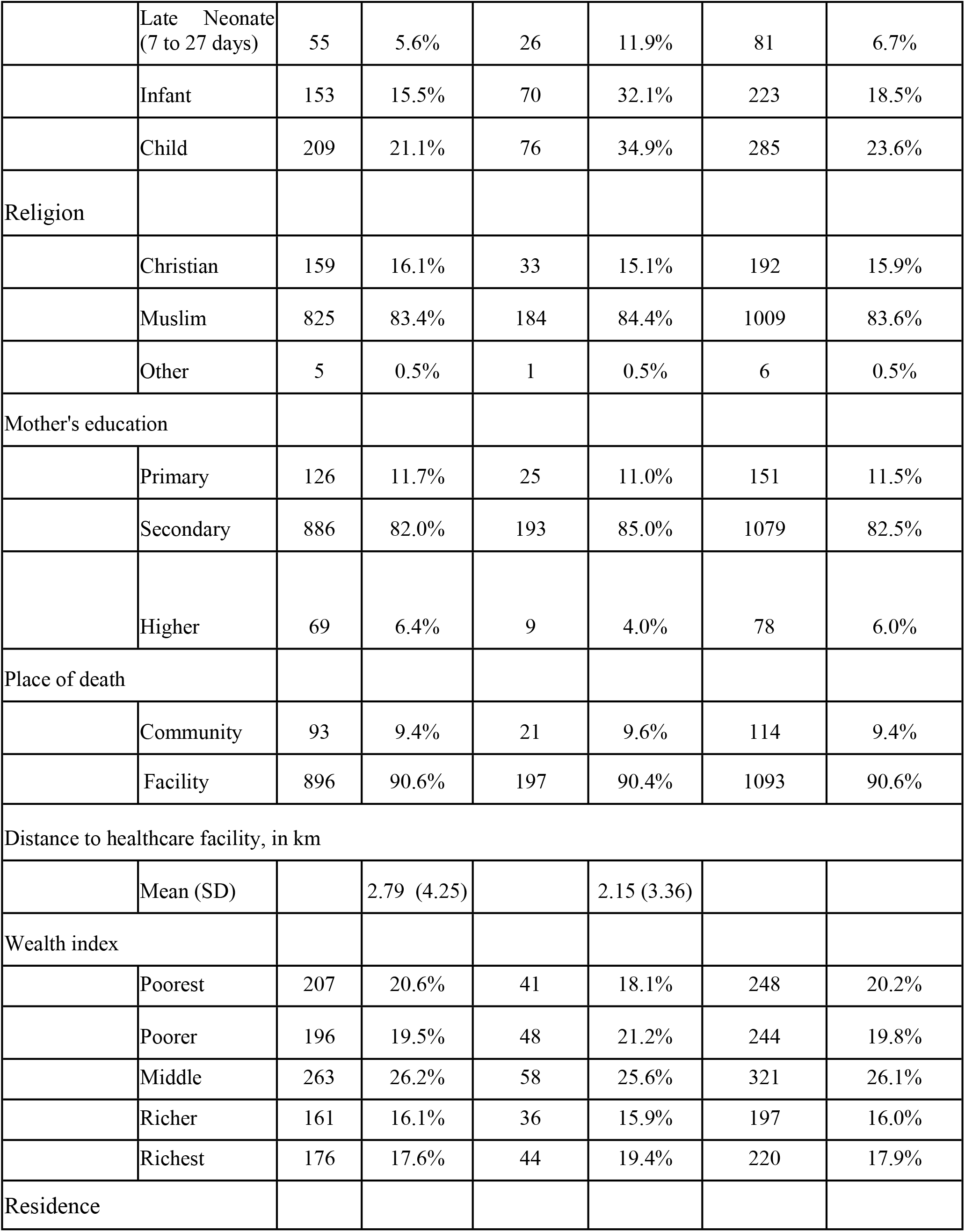
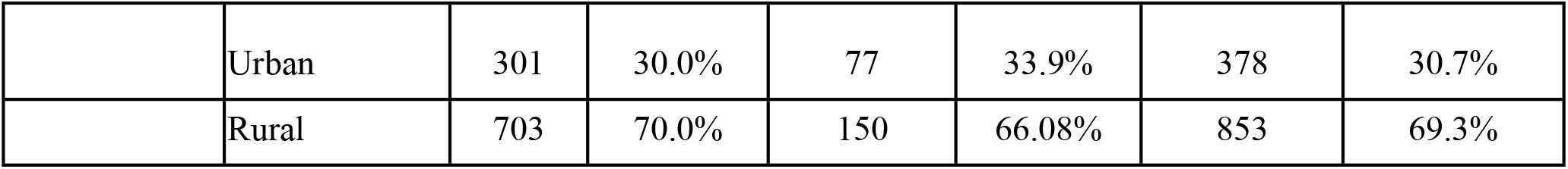
Socioeconomic characteristics of the study population (*n*=1230)

Most deaths occurred in health facilities (90.6%), with little difference by outcome status. The mean distance to the nearest health facility was shorter among children with HIV-related deaths (2.15 km, SD 3.36) compared with those without the outcome (2.79 km, SD 4.25).

Regarding socio-economic status, wealth quintiles were relatively evenly distributed across outcome groups. The poorest quintile represented 18.1% of children with HIV-related deaths, while the richest quintile represented 19.4%. Maternal education was predominantly secondary (82.5%) across the sample, with a smaller proportion of mothers reporting higher education (6.0%). Among children with HIV-related deaths, 4.0% had mothers with higher education, compared with 6.4% among those without the outcome.

#### 3.1.2 Wealth distribution and absence of a socioeconomic gradient

Fig 1 illustrates the mean proportion of HIV-related deaths across wealth quintiles. No clear monotonic gradient is observed. While small differences appear between quintiles, the pattern does not suggest a consistent increase or decrease in risk across the wealth distribution. This visual finding aligns with the descriptive results in Table 1.

**Fig 1.**
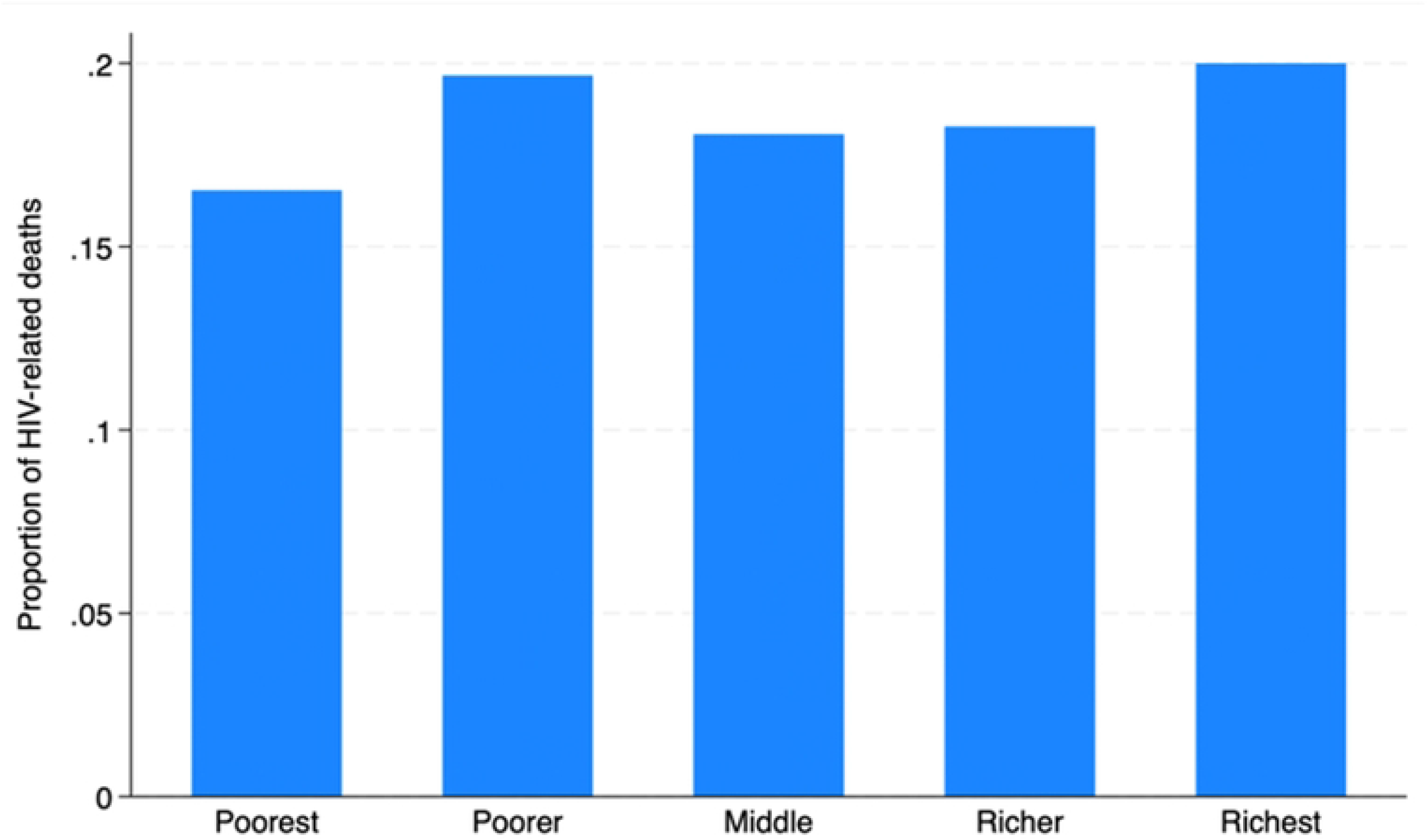
Proportion of HIV-Related Deaths among deceased children under five by household wealth quintile.

### 3.2 Multivariable regression analysis

Table 2 presents the results of stepwise binary logistic regression models examining the association between socio-economic status, health system access, and HIV-related death. Likelihood ratio tests demonstrated that while Model 1 was not statistically significant (χ^2^ = 1.24, p = 0.871), sequential model adjustments significantly improved overall fit in Model 2 (χ^2^ = 23.79,p = 0.005) and Model 3 (χ^2^ = 117.62, p < 0.001).

**Table 2.**
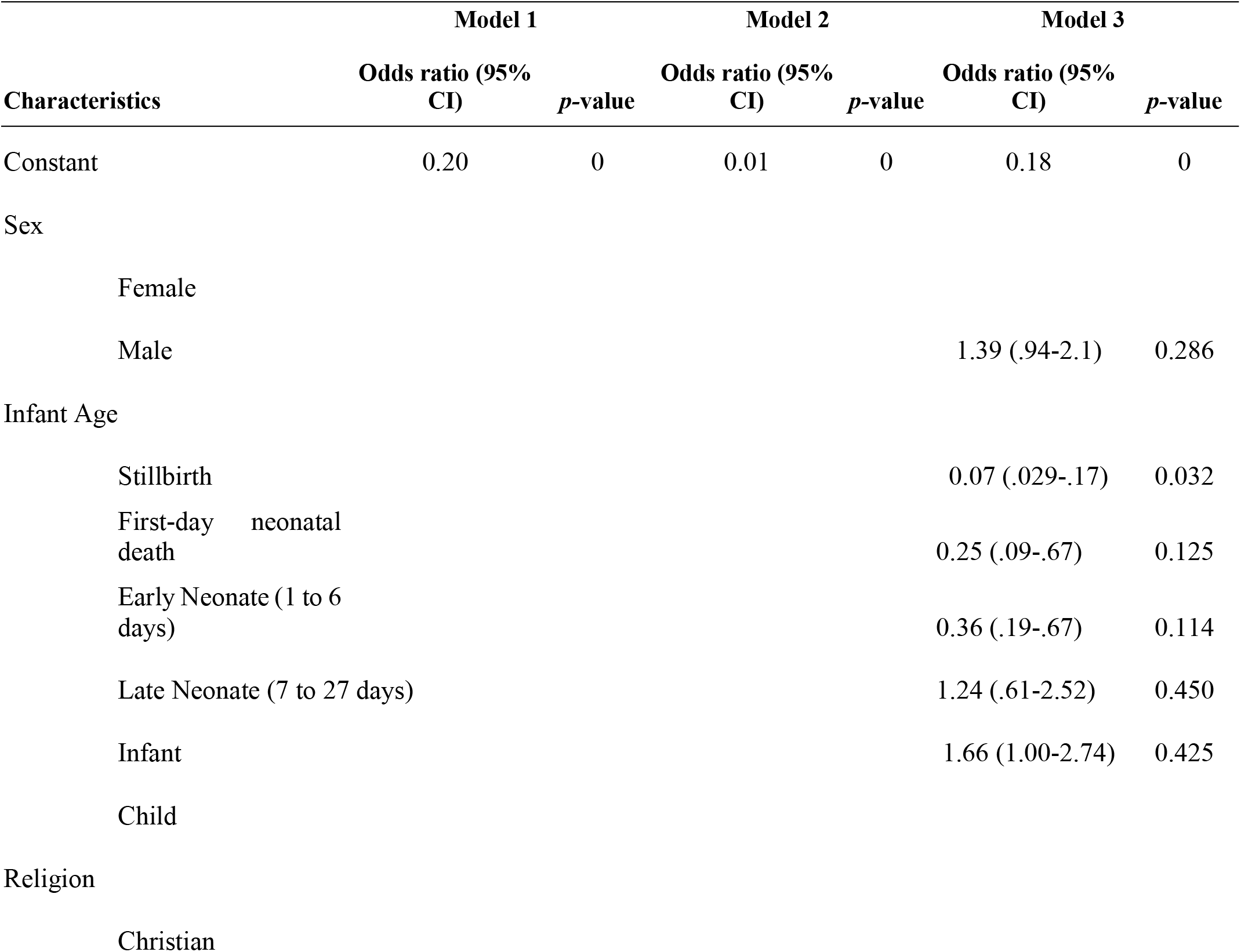
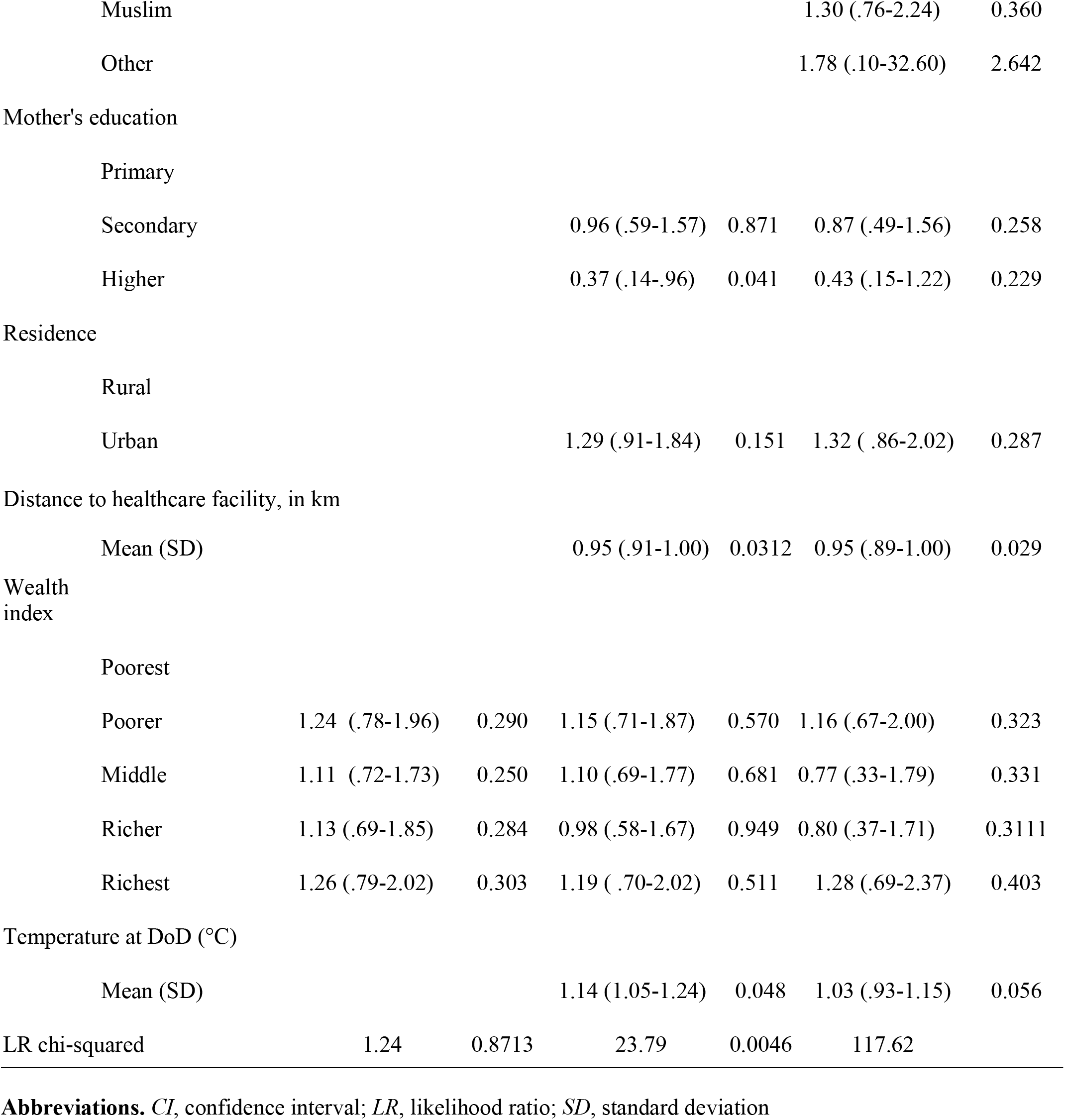
Associations of socioeconomic, maternal, and health system access factors with HIV- related mortality among deceased children under five years of age in Sierra Leone.

|  | **Model 1** |  | **Model 2** |  | **Model 3** |  |
|---|---|---|---|---|---|---|
| **Characteristics** | **Odds ratio (95% CI)** | ***p*-value** | **Odds ratio (95% CI)** | ***p*-value** | **Odds ratio (95% CI)** | ***p*-value** |
| Constant | 0.20 | 0 | 0.01 | 0 | 0.18 | 0 |
| Sex |  |  |  |  |  |  |
| Female |  |  |  |  |  |  |
| Male |  |  |  |  | 1.39 (.94-2.1) | 0.286 |
| Infant Age |  |  |  |  |  |  |
| Stillbirth |  |  |  |  | 0.07 (.029-.17) | 0.032 |
| First-day neonatal death |  |  |  |  | 0.25 (.09-.67) | 0.125 |
| Early Neonate (1 to 6 days) |  |  |  |  | 0.36 (.19-.67) | 0.114 |
| Late Neonate (7 to 27 days) |  |  |  |  | 1.24 (.61-2.52) | 0.450 |
| Infant |  |  |  |  | 1.66 (1.00-2.74) | 0.425 |
| Child |  |  |  |  |  |  |
| Religion |  |  |  |  |  |  |
| Christian |  |  |  |  |  |  |
| Muslim |  |  |  |  | 1.30 (.76-2.24) | 0.360 |
| Other |  |  |  |  | 1.78 (.10-32.60) | 2.642 |
| Mother's education |  |  |  |  |  |  |
| Primary |  |  |  |  |  |  |
| Secondary |  |  | 0.96 (.59-1.57) | 0.871 | 0.87 (.49-1.56) | 0.258 |
| Higher |  |  | 0.37 (.14-.96) | 0.041 | 0.43 (.15-1.22) | 0.229 |
| Residence |  |  |  |  |  |  |
| Rural |  |  |  |  |  |  |
| Urban |  |  | 1.29 (.91-1.84) | 0.151 | 1.32 ( .86-2.02) | 0.287 |
| Distance to healthcare facility, in km |  |  |  |  |  |  |
| Mean (SD) |  |  | 0.95 (.91-1.00) | 0.0312 | 0.95 (.89-1.00) | 0.029 |
| Wealth index |  |  |  |  |  |  |
| Poorest |  |  |  |  |  |  |
| Poorer | 1.24 (.78-1.96) | 0.290 | 1.15 (.71-1.87) | 0.570 | 1.16 (.67-2.00) | 0.323 |
| Middle | 1.11 (.72-1.73) | 0.250 | 1.10 (.69-1.77) | 0.681 | 0.77 (.33-1.79) | 0.331 |
| Richer | 1.13 (.69-1.85) | 0.284 | 0.98 (.58-1.67) | 0.949 | 0.80 (.37-1.71) | 0.3111 |
| Richest | 1.26 (.79-2.02) | 0.303 | 1.19 ( .70-2.02) | 0.511 | 1.28 (.69-2.37) | 0.403 |
| Temperature at DoD (°C) |  |  |  |  |  |  |
| Mean (SD) |  |  | 1.14 (1.05-1.24) | 0.048 | 1.03 (.93-1.15) | 0.056 |
| LR chi-squared | 1.24 | 0.8713 | 23.79 | 0.0046 | 117.62 |  |

**Abbreviations.** *CI*, confidence interval; *LR*, likelihood ratio; *SD*, standard deviation

In Model 1, none of the wealth categories is significantly associated with the outcome, confirming the absence of a direct wealth gradient in HIV-related death. When adjusting for child-level, maternal-level, and health system covariates, Model 2 suggests a protective effect for maternal education. Children of mothers with secondary education have approximately a 4% lower probability of experiencing an HIV-related death compared with those whose mothers have primary education, while higher maternal education is associated with an estimated 63% reduction in the odds of the outcome.

Geographic access was also associated with the outcome. Based on the model estimates, farther distance to the nearest healthcare facility was associated with lower odds of an HIV-related death, with each additional kilometre corresponding to an approximate 4.8% reduction in the odds of the outcome. Temperature on the date of death is also positively associated with the outcome in this model. For each one-degree Celsius increase on the date of death, there is a 14% increase in the odds of HIV-related death of 14%.

Lastly, after running model 3 to further adjust for child age group and sex, we found that child age is associated with the outcome. Compared with older children, late neonates and infants show higher odds of HIV-related deaths, with risk increasing as children survive longer after birth. After adjustment for age and sex, the protective effect of maternal education attenuates and loses statistical significance, suggesting partial mediation by child survival and age at presentation rather than a reversal of direction.

### 3.2 Qualitative results

Analysis of 33 key informant interviews generated interconnected themes describing how PMTCT services are understood, delivered, and constrained within the health system in Sierra Leone. Participants included frontline health workers and health system actors involved in PMTCT service delivery, most affiliated with public-sector institutions, with a small number working in private health facilities. Participant characteristics are summarised in Table 3.

**Table 3:** Characteristics of qualitative study participants.

| Participant characteristics | N (%) |
|---|---|
| Participant category |  |
| Frontline health workers and stakeholders | 33 (100) |
| Professional cadre |  |
| Doctors/ clinical officers | 9 (27.3) |
| Nurses | 5 (15.2) |
| Midwives | 4 (12.1) |
| Pharmacists | 2 (6.1) |
| Community Health workers | 2 ( 6.1) |
| Programme / Global Fund representatives | 2 (6.1) |
| Heads of HIV organisations | 2 (6.1) |
| Director of procurement | 1 (3.0) |
| Clinical, coordination, and health systems staff† | 7 (21.2) |
| Type of Institution |  |
| Public | 30 (90.9) |
| Private | 3 (9.1) |
| Sex |  |
| Female | 12 (36.4) |
| Male | 21 (63.6) |
| Age mean (years) |  |
| Mean (SD)* | 44.2 (8.9) |
| Range | 27-62 |

†Clinical coordination staff: physician officers, regional coordinator, infectious disease physician.
‡Health systems support staff: programme coordinator, health products manager, one unspecified support role.
§Sex inferred from names.
\*Age data available for 18 of 33 participants.
SD = standard deviation.

These themes were organised into two higher-level categories that reflect different layers of influence shaping PMTCT delivery and uptake.

The first category, system-level determinants of PMTCT delivery, captures service availability and access, structural resource constraints, cross-cutting implementation challenges, and donor-driven dynamics that shape planning and service provision. The second category, complex sociocultural and economic barriers to PMTCT uptake and retention, brings together participants’ accounts of stigma and disclosure fears, wider sociocultural norms, and socioeconomic disadvantage that influence women’s engagement with PMTCT services.

Table 4 below presents each theme in detail, supported by illustrative quotations from participants.

**Table 4:** Matrix table. Qualitative findings on prevention of mother-to-child transmission (PMTCT) implementation and barriers.

| Analytical category | Themes | Key findings (Summary) | Illustrative verbatim quotes (KII) |
|---|---|---|---|
| System level determinants of PMTCT delivery | Service availability and access to PMTCT services | PMTCT services are integrated into ANC, delivery, and postnatal care but effective access is undermined by late ANC attendance, women presenting in labor, weak postpartum follow-up, and transport barrier. Many HIV-exposed infants are missed after delivery<br>1) PMTCT integrated across ANC, delivery, postnatal care<br>2) Late ANC, labor presentation, weak follow-up<br>3) Missed HIV-exposed infants after delivery | “Some women came very late for ANC, so detection is delay” (Doctor 5),<br>“Some women only come to facility during labor. They miss the opportunity for early testing and counseling. At that stage intervention options are limited. Early attendance is still a challenge.” (Pharmacist 2).<br>“Low testing of exposed infant is a major challenge.” (Global Fund programme representative 1) |
|  | Structural resource constraints (test kits, ARVs, PCR capacity) | Recurrent stockouts of HIV test kits and ARVs disrupt timely diagnosis and treatment. Limited PCR capacity and delay in results discourage caregiver follow-up and contribute to loss to care<br>1) Stockouts of HIV test kits and ARVs<br>2) Limited PCR capacity and delayed results<br>3) Delayed care and loss to follow-up | “There are times when HIV test kits are not available in the facility. Even if a pregnant woman comes for ANC, we cannot test her immediately. This leads to missed opportunities for early detection. Stockout is a major operational challenge.” (Nurse 3),<br>“When ARVs are not available, women miss doses. Some stop treatment completely. This increases the risk of vertical transmission, drug availability is very important.” (Community Health Officer 2).<br>“Expired kits happen because of improper stock management” (Doctor 4), |
|  | Data, target and donor-driven constraints | PMTCT planning and procurement are shaped by donor target rather than true burden. Poor quality data and report gaps distort quantification and contribute to stockout and misallocation of commodities<br>1) Donor-driven targets over true burden<br>2) Poor data quality and reporting gaps<br>3) Misallocation and stockouts of commodities | “The target is given to us, we don’t always quantify base on real number” (Pharmacist 1).<br>“Well, there are lapses with every system. If the quality of the data is not good, it means the quality of what we do would also not be good. If there are facilities that are not reporting, it means the district is going to get far less than what they need. If the figures they report are higher than what the numbers they actually have on treatment, it means we are going to send more commodities than what is needed.” (Supply chain and quantification manager 1). |
|  | Cross cutting implementation challenges | Human resource shortages, weak tracking systems, limited funding, and poor-follow-up mechanisms undermine continuity of PMTCT care for mother-bay pairs<br>1) Coordination and workflow gaps<br>2) Staff shortages and training needs<br>3) Monitoring and accountability challenges | “Lack of follow up, is always, always. That would be an issue also. Yes, it should always be challenge. Because as I still access to care, it’s a problem. You understand? For all polar things will always be a challenge.” (Doctor 2).<br>“We don’t have waiting list. We don’t have appointments, book and even no waiting list. That’s only two things that we don’t have in our center.” (Nurse 3). |
| Complex sociocultural and economic barriers to PMTCT uptake and retention | Stigma, disclosure fears and sociocultural barriers | Fear of disclosure, partner reactions, marital conflict and community stigma strongly affect testing, ART adherence and retention in PMTCT service. Confidentiality concerns are especially acute in small communities<br>1) Fear of disclosure and partner reactions<br>3) Stigma and sociocultural barriers<br>3) Confidentiality concerns in small communities | “Many women fear disclosing their HIV status to their partners. They are afraid of blame or violence. Because of this, some hide their drugs. This affects adherence.” (Doctor 3).<br>“Women avoid nearby facility because they fear being recognized.” (Midwife 2),<br>“Some patients don’t want to identify themselves when collecting ART.” (Pharmacist 2). |
|  | Wider structural norms | Gendered power relation exploitation around marriage and motherhood, and community level stigma shape women’s ability to engage consistently with PMTCT services. Fear of marital instability, blame and social exclusion influences decisions about testing, disclosure and continuation of care. These norms reinforce silence around HIV and constrain autonomous health seeking behavior<br>1) Gender power dynamics and social norms<br>2) Fear of marital instability and social exclusion<br>3) Limited autonomy in health-seeking behavior | “Fear of marital conflict makes women hide their status” (Global Fund representative 1)<br>“Community stigma is still very strong. Women fear being at HIV service points. Some travel far to seek care” (Doctor 2) |
|  | Socio economic disadvantage and practical barriers to care | Poverty, distance and transport costs frequently interrupt follow-up<br>1) Poverty and financial constraints<br>2) Distance and transport barriers<br>3) Interrupted follow-up | “Distance is a big challenge; they stay far and missed appointments.” (Doctor 5).<br>“The financial aspect. Because for the long distance, as long as she’s depressed or she have medical problems, she’ll look for, she’ll come to the facility no matter what, how the distance is. But the financial constraints are a challenge.” (Midwife 3). |

#### System-level determinants of prevention of mother-to-child transmission delivery

##### Service availability and access to PMTCT services

Although PMTCT services were described as formally integrated across antenatal, delivery, and postnatal care, participants consistently emphasized that system-level barriers disrupt the continuum of care, beginning with delayed entry into services and extending into weak postnatal follow-up.

The starting point of this sequence was delayed or absent engagement with antenatal care. Providers explained that many women do not access antenatal health services or attend routine antenatal visits early in pregnancy, limiting opportunities for timely HIV testing and ART initiation. This delayed entry was attributed to a combination of maternal attitudes toward care and late HIV diagnosis, as summarized by one key informant:

> *“Negative attitude towards testing contributing to transmission.” (Pharmacist 1)*

When women either do not present early for care or decline HIV testing during early visits, HIV infection is identified late, narrowing the window for effective PMTCT interventions. Several respondents emphasized that antenatal care is often initiated late in pregnancy:

> *“Some women came very late for ANC, so detection is delay.” (Doctor 5)*

A district-level participant highlighted the programmatic implications of late booking:

> *“Many women attend ANC very late in pregnancy. Some come when they are already seven or eight months pregnant. This delays HIV testing and treatment. It increases the risk of transmission.” (HIV programme director 1)*

For some women, delayed care-seeking progresses further into no antenatal contact at all, with first presentation occurring during labor. Participants noted that at this stage, PMTCT options are extremely limited:

> *“Some women only come to facility during labor. They miss the opportunity for early testing and counseling. At that stage intervention options are limited. Early attendance is still a challenge.” (Pharmacist 2)*

Providers linked this pattern to limited understanding of the purpose of antenatal care, explaining that some women perceive ANC as necessary only in the presence of illness, further delaying HIV detection:

> *“Lack of awareness contributes to late ANC booking. Some women think ANC is only necessary when sick. This belief affects early HIV detection. Community education is needed.” (Nurse 6)*

Delays in diagnosis and treatment initiation then extend into the postnatal period, where gaps in continuity of care become apparent. Participants described frequent loss to follow-up after delivery, with many mothers failing to return with their infants for postnatal services and early infant diagnosis:

> *“After discharge, mother doesn’t always return with the babies.” (Nurse 2)*

As a result, testing among HIV-exposed infants remains low:

> *“Low testing of exposed infant is a major challenge.” (Global Fund programme representative 1)*

Once postnatal visits are missed, providers reported limited system capacity to trace and re-engage mothers and infants, particularly in the context of fear of stigma or relocation:

> *“After delivery some mothers do not return with the babies for testing. They may be relocate or fear stigma. Once they miss the first appointment, tracking becomes difficult. Infant testing coverage is affected.” (Community Health Officer 1)*

These findings indicate a progressive chain of system-level constraints. Limited awareness and attitudes toward care contribute to delayed or absent antenatal attendance, leading to late HIV diagnosis and reduced effectiveness of PMTCT interventions. Presentation for the first time during labor further narrows available options, while weak continuity between delivery and postnatal services results in poor follow-up of HIV-exposed infants and low infant testing. Although PMTCT services are formally available across antenatal, delivery, and postnatal care, breakdowns at successive points in the care pathway accumulate, limiting effective access for many women and their infants.

##### Structural resource constraints (test kits, ARVs, PCR capacity)

Providers repeatedly emphasized structural resource constraints as a major challenge undermining PMTCT implementation. These constraints included recurrent stockouts of HIV test kits, limited laboratory capacity for PCR, irregular supply of ARVs and prophylaxis, and weaknesses in stock management at both facility and central levels.

Participants linked diagnostic stockouts and PCR limitations to broader supply chain weaknesses at central and facility levels. A district or central-level respondent explained that:

> *“Stock out at central level affects all facilities.” (Pharmacist 1)*

Delays in distribution were also described as contributing directly to facility-level stockouts:

> *“When there are delays in distribution they may likely result in stock out at facility level.” (Supply chain and quantification manager 1)*

Another respondent explicitly connected shortages to reporting and forecasting problems:

> *“Stock outs sometimes happen because facilities do not submit report on time. Poor reporting affects quantification and forecasting. This leads to shortages at facility level. It is a system issue.” (Midwife 1)*

In addition to outright shortages, respondents described weaknesses in stock management that contribute to wastage:

> *“Expired kits happen because of improper stock management.” (Doctor 4)*

These system-level weaknesses were experienced most directly at the facility level. Several informants described periods where essential HIV test kits were unavailable, forcing staff to turn women away or ask them to return later. One provider noted:

> *“There are times when HIV test kits are not available in the facility. Even if a pregnant woman comes for ANC, we cannot test her immediately. This leads to missed opportunities for early detection. Stockout is a major operational challenge.” (Nurse 3)*

Another respondent reported similar experiences:

> *“There are periods when HIV and test kits are not available at facility. Pregnant women come but cannot be tested. We ask them to return later, some never come back.” (Infectious disease physician 1)*

Participants emphasized that these missed opportunities have direct consequences for early HIV diagnosis and timely initiation of ART, particularly for women who already present late in pregnancy.

Laboratory capacity, particularly access to PCR for early infant diagnosis, was described as another major diagnostic constraint. Several respondents explained:

> *“Without PCR early infant’s diagnosis is difficult.” (Physician Officer 1)*

Delayed PCR results were also reported to affect follow-up of HIV-exposed infants:

> *“Delay PCR results discourage caregiver, when result take too long they lose interest, some stop coming completely.” (Doctor 4)*

After diagnosis, providers described challenges related to inconsistent availability of ARVs and prophylaxis, which interrupt treatment continuity for pregnant women and mothers. One clinician observed:

> *“When ARVs are not available, women miss doses. Some stop treatment completely. This increases the risk of vertical transmission, drug availability is very important.” (Community Health officer 2)*

Another explained that facilities sometimes resorted to informal coping strategies:

> *“Sometimes we borrow drugs from nearby facilities.” (Pharmacist 1)*

A few participants acknowledged that stock availability had improved compared to previous years. However, they continued to describe PMTCT commodity supply as inconsistent, with ongoing disruptions affecting HIV testing, infant diagnosis, and treatment continuity.

##### Cross-cutting implementation challenges

Participants described a range of broader implementation challenges that undermine PMTCT service delivery across all levels of the health system. These challenges cut across facility, district, and national levels and included human resource constraints, high workload, weak referral and follow-up systems, limited integration of services, inadequate mechanisms to track HIV-positive mothers and exposed infants, and persistent gaps in funding and materials.

Front-line providers frequently highlighted human resource shortages and the absence of basic systems for scheduling and tracking clients:

> *“We don’t have waiting list. We don’t have appointments, book and even no waiting list. That’s only two things that we don’t have in our center.” (Nurse 3)*

This absence of basic organizational systems was perceived as both a symptom and a cause of chronic overload, with direct implications for continuity of care. Weak follow-up was repeatedly described as a persistent challenge within this context:

> *“Lack of follow up is always a challenge.” (Midwife 3)*

Participants at policy and program levels highlighted weak tracking systems and poor linkage between facilities as structural barriers to effective follow-up. These challenges were closely tied to data quality and reporting gaps:

> *“If the quality of the data is not good, the quality of what we do would also not be good.” (Supply chain and quantification manager 1)*

Others emphasized the need for additional manpower and sustained external support to strengthen PMTCT delivery and address persistent system gaps. In this context, *sensitization* referred to community outreach and information activities intended to support PMTCT service delivery and follow-up:

> *“There is always room for support, always room for support, especially as we see to cut the chain of transmission. More sensitization, more materials. The diagnostics. We need to do our, what I say to pinpoint the patient that are taking and other things, more manpower needs to be emphasized. If those things are there, especially sensitization, because there are stigma towards this HIV, especially mother to child area. The manpower, the support we need.” (Doctor 1)*

Gaps in staffing, scheduling, client tracking, data systems, and operational capacity undermine continuity of care across the PMTCT cascade.

##### Target and donor-driven constraints

Another cross-cutting system issue relates to how donor targets and funding structures shape PMTCT delivery. Participants described an environment where Global Fund and other donor targets strongly influence planning and resource allocation, but are often misaligned with local epidemiology and service realities.

Several informants explained that quantification is frequently driven from above:

> *“The target is given to us, we don’t always quantify based on real number.” (Pharmacist 1)*

Central-level respondents linked national HIV estimates, donor targets, and procurement decisions:

> *“For example, UNAIDS will say Sierra Leone, you have 80,000 people living with HIV. The Global Fund targets will maybe be 70,000.” (Supply chain and quantification manager 1)*

This structure effectively caps procurement and planning, irrespective of unmet need. Limited domestic funding reinforced donor dependence:

> *“We have targets in the National Strategic plan, but we cannot buy those targets because we don’t have the funds.” (Supply chain and quantification manager 1)*

These accounts illustrate how donor dependence and target-driven planning constrain PMTCT delivery, limiting the system’s ability to respond flexibly to ongoing vertical transmission risk.

#### Complex sociocultural and economic barriers to PMTCT uptake and retention

##### Stigma, disclosure fears, and sociocultural barriers

Stigma, fear of disclosure, and wider sociocultural norms emerged as powerful and persistent barriers to PMTCT uptake, adherence, and retention. Participants consistently described how women’s fear of partner reactions, marital conflict, and community judgement undermines both HIV testing and sustained engagement in care.

Providers reported that many women were reluctant to disclose their HIV status to their partners because they feared blame, abandonment, or violence. One respondent explained that:

> *“Many women fear disclosing their HIV status to their partners. They are afraid of blame or violence. Because of this, some hide their drugs. This affects adherence.” (Doctor 3)*

Another health worker observed that:

> “Some women stop coming because they are afraid their husband will know.” (Regional health coordinator 1)

and yet another added that:

> *“Fear of marital conflict makes women hide their status.” (Global Fund representative 1)*

These accounts illustrate how gendered power relations and fear of intimate partner conflict shape women’s ability to attend services consistently and adhere to ART, even when PMTCT services are technically available.

Within this broader context of stigma, concerns about confidentiality emerged as a key mechanism through which stigma operates, particularly in small and close-knit communities. Providers explained that women often avoid nearby facilities because of fear of recognition by neighbours, relatives, or acquaintances. A midwife noted that:

> *“Women avoid nearby facility because they fear of been recognize.” (Midwife 2)*

Another respondent remarked that:

> *“Confidentiality is a concern especially in a small community.” (Nurse 5)*

These fears extended into facility interactions themselves. A pharmacist explained that:

> *“Some patients don’t want to identify themselves when collecting ART.” (Pharmacist 2)*

A nurse working in a referral hospital described how women explicitly request that certain staff not be involved in their care because of social proximity:

> *“Like for some of them they find out that there is a close friend to her in that facility or there is a family. Somebody that is a part of their family is in that facility. That one cause a stigma to this woman. Sometimes they will tell us that ‘No, please, even this nurse, I don’t want her to know my status’, but this nurse is working in the facility. How can you cover unless the nurse has to look, but it’s a secret, it is confidential. So I normally tell them that this one is confidential.” (Midwife 3)*

Participants emphasized that these confidentiality concerns are not isolated incidents but are embedded in broader community stigma. One respondent explained that:

> *“Community stigma is still very strong. Women fear being at HIV service points. Some travel far to seek care. This affects continuity of care.” (Doctor 2)*

Another highlighted how even rumours can undermine service use:

> *“Confidentiality is a major concern for clients. They worry health workers might disclose their status. Even rumors discourage service use. Trust is very important.” (Midwife 3)*

Providers further linked stigma and fear of disclosure to psychological distress following diagnosis. One respondent noted that:

> *“Psychological stress affects many HIV-positive mothers. Some become depressed and withdrew from care. Without psychosocial support, adherence becomes difficult. Mental health services are limited.” (Nurse 5)*

Another health worker described how difficulty accepting an HIV diagnosis often leads to disengagement from care, even when support is offered:

> *“The challenges here are for me, before this time we were having challenges of supplies. But now it’s better. And we are having challenges of those mothers to accept their status sometimes whenever I say to give them to care. Even if you take them along to the facility, they’ll not continue.” (Midwife 3)*

This pattern was echoed by another participant, who observed that:

> *“After testing positive some become depressed and stop coming.” (Global Fund representative 1)*

Across participants’ accounts, stigma and disclosure fears appear as a persistent thread shaping engagement with PMTCT services, influencing testing uptake, continuity of ART, choice of facility, postnatal attendance, and infant testing.

##### Socio-economic disadvantage and practical barriers to care

Closely intertwined with stigma are the socio-economic and practical barriers that shape women’s ability to engage consistently with PMTCT services. Participants repeatedly highlighted long distances to health facilities as a major structural barrier, particularly for routine antenatal visits, postnatal follow-up, and infant testing.

Distance to health facilities was frequently described as discouraging early attendance and continuity of care. One midwife explained that:

> *“Distance is a big challenge, they stay far and missed appointment.” (Doctor 5)*

Another respondent linked distance directly to delayed care-seeking:

> *“Distance to the health facility discourages early ANC attendance. Transport cost is also a problem. Women delay until they have complications, this affects PMTCT outcome.” (Midwife 4)*

While distance was identified as the primary access barrier, participants emphasized that its impact is largely mediated through transport costs. Even when women understand the importance of follow-up, the cost of reaching facilities repeatedly was described as prohibitive. One respondent noted that:

> *“Transport cost affects follow-up visit.” (Doctor 2)*

Several respondents emphasized that transport expenses pose a recurrent challenge, especially given the need for monthly drug refills and multiple follow-up visits. One provider stated that:

> *“Some participants cannot afford transport to come every month for drug.” (Pharmacist 1)*

Another nurse reflected on how financial constraints shape routine care-seeking differently from emergency care:

> *“The financial aspect is a problem. Because for the long distance, as long as she’s depressed or she have medical problems, she’ll look for, she’ll come to the facility no matter what, how the distance is. But the financial constraints are a challenge.” (Midwife 3)*

These accounts illustrate how distance, transport costs, and material deprivation operate together. Distance creates the initial barrier, transport costs determine whether women can overcome it, and wider poverty constrains their ability to prioritise repeated clinic visits over competing household needs.

These practical constraints repeatedly interrupted ART pick-up and infant follow-up, reinforcing gaps in PMTCT engagement even when services were formally available.

## 4. Discussion

### 4.1 Summary of main findings

This study sought to examine the extent to which socioeconomic position and health system access are associated with HIV-related child mortality among deceased children under five years of age in Sierra Leone, and to use qualitative evidence to explain observed quantitative patterns. The quantitative analysis showed no evidence of a socioeconomic gradient in HIV-related deaths by household wealth. Instead, distance to healthcare facilities, child age and maternal education were associated with the outcome in the multivariable models. Higher maternal education showed a protective association in partially adjusted models, though this effect attenuated after adjustment for child age and sex. The qualitative findings help explain how maternal education may influence PMTCT outcomes indirectly. The mixed-methods approach provided insights beyond standalone quantitative analysis. While the quantitative analysis identified associations between maternal education, distance to facilities and HIV-related child mortality, the qualitative findings provided the operational mechanism underpinning these associations. By capturing health system breakdowns such as delayed antenatal care entry, supply chain disruptions, and postpartum retention gaps, the qualitative data provided crucial context for how structural vulnerabilities translate into missed PMTCT opportunities across Sierra Leone.

### 4.2 Explanation of findings and comparison with existing literature

The lack of a significant association between household wealth and HIV-related mortality in our sample differs from prior evidence linking lower socioeconomic status to higher HIV-related mortality [15]. It also contrasts with findings from Mozambique showing that HIV infection is disproportionately concentrated among poorer populations [17]. However, direct comparison with these studies requires caution due to differences in outcome measures. While the prior literature focused on general HIV acquisition risk [15] or adult/population-level HIV infection prevalence [17], our study specifically evaluated a composite outcome of HIV-related mortality among deceased children under five. Consequently, socio-economic disparities may manifest differently when predicting infection acquisition versus acute pediatric mortality. On the other hand, patterns reported in Sierra Leone suggest greater HIV vulnerability among women in middle, richer, and richest wealth quintiles compared with those in the poorest households [18], a pattern that contradicts the absence of a clear poverty gradient observed in our analysis. At the same time, the SES–HIV relationship is not uniform across settings; for example, in Uganda, higher household wealth has been associated with increased HIV vulnerability [16]. In this post-mortem surveillance context, however, wealth may be less discriminating than structural access and health system factors that affect families across the socioeconomic spectrum. The quantitative finding that distance to health facilities is associated with reporting an HIV-related death supports this interpretation. Qualitative accounts indicate that geographic distance, combined with transport costs and weak follow-up systems, delays antenatal attendance, limits continuity of care, and reduces opportunities for timely HIV testing, ART initiation, and early infant diagnosis. Similar patterns have been reported in other low-resource settings, where barriers such as service availability, supply reliability, and geographic access contribute to gaps in PMTCT uptake and continuity of care, alongside other patient and health-system constraints [5,6].

The analysis revealed an association between distance to the nearest health facility and recorded HIV-related mortality, with greater distance paradoxically linked to lower odds of a documented HIV-related outcome. Although follow-up data were limited, loss to follow-up represents a plausible mechanism, consistent with observations elsewhere [35]. Patients who are unable to attend scheduled appointments are rarely traced by health systems, and if death occurs at home, it often remains unlinked to their HIV status in official records. Although no study to date has formally modeled geographic distance to health facilities as a predictor of HIV death under-reporting within a regression framework, related evidence supports this hypothesis. De Paula and colleagues demonstrated that dying outside a hospital increased the odds of HIV being omitted from the death certificate by 2.6-fold in Brazil (OR = 2.61, 95% CI: 2.31–2.95). This finding suggests that distance (by increasing the likelihood of out-of-facility death) may contribute to the under-reporting of HIV-related mortality [36].

The qualitative findings help explain this pattern by describing how geographic barriers shape engagement with PMTCT services across the care pathway. Health workers consistently reported that long distances and transport costs discourage early antenatal attendance and repeated facility visits, leading to delayed HIV testing and treatment initiation. They further explained that these barriers become more pronounced after delivery, when continuity of care depends on mothers returning with their infants for postnatal follow-up and early infant diagnosis. According to participants, many mothers do not return after discharge, and once postnatal appointments are missed, facilities often lack the staffing, scheduling systems, and tracking capacity needed to trace and re-engage mothers and HIV-exposed infants. In more remote and rural settings, these challenges are intensified, increasing the likelihood that infants are never tested and that deaths occur outside the formal health system. As a result, HIV may be less likely to be diagnosed and less likely to record HIV-related death among children living farther from health facilities, even when underlying risk remains high.

Maternal education showed a protective association in the expected direction, with higher education linked to lower odds of HIV-related death, although this effect attenuated after adjustment for child age. This pattern is consistent with prior evidence demonstrating educational gradients in HIV vulnerability, with lower educational attainment associated with higher HIV risk and reduced access to prevention and treatment services, which are proxies for HIV mortality [16,17]. Qualitative evidence indicates that while higher educational attainment may facilitate engagement with PMTCT services, it does not fully mitigate the effects of stigma, fear of disclosure, confidentiality concerns, and psychological distress that contribute to disengagement [12,13]. Health system constraints, including stockouts of HIV test kits and antiretrovirals and broader weaknesses in laboratory capacity, combined with fragmented and donor-dependent planning, can limit the effectiveness of PMTCT services even when women present for care [5]. Taken collectively, these findings are consistent with evidence that PMTCT outcomes are shaped not only by individual engagement but also by consistent availability of HIV testing and treatment services and by social conditions that influence women’s ability to access and remain in care [2,3].

### 4.3 Limitations and Strengths

This study has several limitations. The quantitative analysis is based on deceased children enrolled in CHAMPS surveillance, and missing HIV status and cause-of-death data led to exclusions that may limit generalizability and introduce selection bias. To partially address this limitation, we expanded the outcome definition to include intermediate conditions, including pneumonia and sepsis, which are common opportunistic and HIV-associated causes of morbidity and mortality in immunocompromised children and may reflect underlying or undiagnosed HIV infection. However, some degree of selection bias likely remains. Although extensive diagnostic procedures enhance outcome classification, missing data led to exclusions that may introduce selection bias. Religious and cultural factors may also have influenced participation in post-mortem surveillance. According to CHAMPS data management teams, families from Muslim backgrounds may be more likely to decline consent for post-mortem procedures, which could result in underrepresentation and potential bias. Qualitative findings reflect the perspectives of health workers and program stakeholders and may not fully capture caregivers’ lived experiences. The counterintuitive association observed between geographic distance to healthcare facilities and HIV-related mortality should be interpreted with caution and likely reflects differences in access to care and case detection rather than a true protective effect of distance; children living closer to healthcare facilities may be more likely to access services, undergo HIV testing, and be identified within the surveillance system, while those residing farther away may experience delayed care-seeking, reduced diagnostic coverage, or die outside formal healthcare settings, resulting in under-detection of HIV-related outcomes. In addition, residual confounding may persist if populations living farther from facilities differ systematically in age, health status, or risk profiles, although adjustment for age in model 3 did not alter the direction of the association. Selection bias may also arise if sicker children are more likely to reside closer to healthcare facilities, and the relationship between distance and mortality may be non-linear across its full range, such that the assumption of linearity in the logit model could contribute to unexpected estimates; alternative specifications such as polynomial terms may be more appropriate. Finally, reverse causality or other unmeasured factors may influence both geographic proximity to care and mortality outcomes.

### 4.4 Conclusions and future directions

This study shows that HIV-related deaths among children in Sierra Leone might be shaped less by household wealth than by structural access barriers, health system constraints, and sociocultural factors that undermine PMTCT effectiveness. Improving geographic access, strengthening supply chains and laboratory capacity, enhancing follow-up systems, and addressing stigma and confidentiality concerns are critical for reducing vertical transmission. Future research should disentangle the unmeasured factors underlying the observed association between geographic access and HIV-related mortality, including how deaths are identified through routine healthcare contact, detected post hoc through CHAMPS surveillance, or remain undetected, and how these pathways of detection vary with distance to health facilities. Additional work is also needed to incorporate caregiver perspectives, explore community-based follow-up models, and assess interventions that integrate psychosocial support with PMTCT service delivery to address persistent gaps along the prevention cascade.

## Availability of data and material

The dataset will be available to authors upon reasonable request. Data access may be obtained by submitting a request to the Site Senior Data Manager of The Africa Research Collaborative (The ARC), on behalf of the Child Health and Mortality Prevention Surveillance, Sierra Leone. Requests will be reviewed by the Science Innovation and Research Team of the same organization.

## Ethical Approval

Ethical approval for the use of CHAMPS post-mortem surveillance data was obtained from the Sierra Leone Ethics and Scientific Review Committee and institutional review boards overseeing CHAMPS implementation (SLESRC–002/05/2024). All qualitative participants provided written informed consent. Confidentiality was maintained through de-identification and secure data storage.

## Consent for publication

Not applicable

## Authors Contribution

Supervision. OG, SA, CRR, IUO

Concept and study design. SC, OG, CRR, SA, IUO Data collection/extraction. SC, CRR, DK, HSS Data curation. SC, CRR, DK

Formal analysis. SC, CRR

Review, interpretation and discussion of results. SC, OG, CRR, SA, IUO, DK, HSS

## Acknowledgments

We acknowledge the study participants, notably the catchment communities and families of the deceased U5s who were included in this research. We are grateful to the Social and Behavioral Sciences unit for their invaluable contributions to the qualitative component of this research, as well as to the leadership of the Africa Research Collaborative (The ARC), on behalf of the Child Health and Mortality Prevention Surveillance (CHAMPS) in Sierra Leone. We acknowledge Prof Dan Smith for his selfless commitment and the financial support to Sabrina Clumeck through the Watson School Africa Initiative, Brown University, USA. The research findings and conclusions are those of the authors and do not necessarily represent the views of the funders.

## Supplementary information captions

Supplementary Text A. CHAMPS Procedure and Data Collection

Supplementary Text B. Measuring proximity of HIV-related mortality cases to healthcare facilities

Supplementary Text C. Qualitative Interview Guides

